# Prevalence of excess adiposity and clinical obesity in a Mexican nationally representative survey

**DOI:** 10.64898/2026.08.18.26360751

**Authors:** Mario Cesar Torres-Chavez, Neftali Eduardo Antonio-Villa, Mauricio Gonzalez-Arias, Diego Araiza-Garaygordobil, Pablo Martinez-Amezcua

## Abstract

Body mass index (BMI) alone may underestimate clinically relevant obesity because it does not capture central fat distribution. We compared obesity prevalence in Mexico using BMI-only criteria, adiposity-confirmed criteria, and the clinical obesity definition proposed by the Lancet Diabetes and Endocrinology Commission. We conducted a population-based, cross-sectional study of 13,160 adults aged 18 years or older who participated in the 2018-2019 Mexican National Health and Nutrition Survey (ENSANUT). Obesity prevalence was estimated through survey-weighted analyses that accounted for the complex sampling design. The weighted prevalence of obesity based on BMI was 34.5% (95% CI, 33.1-35.9), while 30.9% (95% CI, 29.6-32.2) met criteria for clinical obesity. One quarter of individuals with clinical obesity had a BMI under 30 kg/m^2^, a phenotype more common among older adults. Half of adults with a BMI under 30 kg/m^2^ showed elevated central adiposity. BMI alone underestimates clinically relevant obesity in Mexican adults. Adding waist-based measurements could improve the identification of individuals with excess fat and metabolic risk, both in clinical settings and population monitoring.

## INTRODUCTION

Body mass index (BMI) is widely used to define obesity for clinical and surveillance purposes, yet it does not distinguish adiposity from lean mass nor capture central fat accumulation, a key cardiometabolic risk factor. As a result, reliance on BMI alone may lead to under-identification of individuals at risk for obesity-related cardiometabolic complications.(1) To address these limitations, the Lancet Diabetes & Endocrinology Commission proposed a clinical framework that conceptualizes obesity as a disease characterized by confirmed excess adiposity—assessed by direct imaging methods or indirect anthropometric proxies—alongside evidence of obesity-related metabolic dysfunction.(2) This framework aims to improve risk stratification and align obesity classification with clinical relevance rather than solely body size.

Despite its potential implications for diagnosis, treatment prioritization, and population surveillance, the impact of this clinical obesity definition on national obesity prevalence has not been quantified in most settings. This gap is particularly relevant for Mexico, a country with one of the highest obesity rates defined by BMI prevalence worldwide and a substantial burden of cardiometabolic disease. (3) In this study, we compared obesity prevalence in Mexico using BMI-only criteria, adiposity-confirmed criteria, and the clinical obesity definition proposed by the Lancet Diabetes & Endocrinology Commission using nationally representative survey data.

## METHODS

We analyzed data from adults aged ≥18 years participating in the 2018–2019 Mexican National Health and Nutrition Survey (ENSANUT), a probabilistic, multistage, stratified survey designed to represent the national population. Detailed survey methods have been published previously.(4)

Obesity was defined as body mass index (BMI) ≥30 kg/m^2^. Following the Lancet Diabetes & Endocrinology Commission framework, excess adiposity was operationalized as any of the following: BMI ≥40 kg/m^2^; BMI ≥30 kg/m^2^ plus at least one elevated anthropometric index (waist circumference ≥102 cm in men or ≥97 cm in women, or waist-to-height ratio ≥0.5); or at least two elevated indices among individuals with BMI <30 kg/m^2^. Clinical obesity additionally required evidence of organ dysfunction, defined as at least one metabolic abnormality: fasting glucose ≥126 mg/dL or HbA1c ≥6.5%, triglycerides ≥200 mg/dL, HDL cholesterol <40 mg/dL, or blood pressure ≥140/90 mmHg. These thresholds were more stringent than those originally proposed by the Commission to improve specificity for clinically relevant metabolic impairment in this population.(2,5,6)

Survey-weighted estimates accounted for the complex sampling design. Weighted prevalences were stratified by sex, age group (18–29, 30–39, 40–49, 50–59, and ≥60 years), educational attainment (none, primary/secondary, high school/some college, college or more), urban or rural residence, and monthly household income quartiles (Q1 ≤800 MXN, Q2 801–1,800 MXN, Q3 1,801–10,000 MXN, and Q4 >10,000 MXN). All analyses were conducted using R software (version 4.5.0).

## RESULTS

Among 13,160 Mexican adults (representing 82,710,347 individuals), the weighted prevalence of obesity defined by BMI was 34.5% (95% CI, 33.1–35.9) (**Table 1**). Estimates of excess adiposity varied depending on the anthropometric definition, ranging from 26.9% when defined by BMI plus waist circumference to 33.3% when defined by BMI plus waist-to-height ratio. Overall, 30.9% of adults met the criteria for clinical obesity. Women had a lower prevalence of obesity based on BMI than men (29.9% vs 38.3%), but had a similar prevalence of clinical obesity (31.1% vs 30.7%). The prevalence of clinical obesity increased with age, from 17.3% among adults aged 18–29 years to 38.6% among those aged 50–59 years.

**Table.**
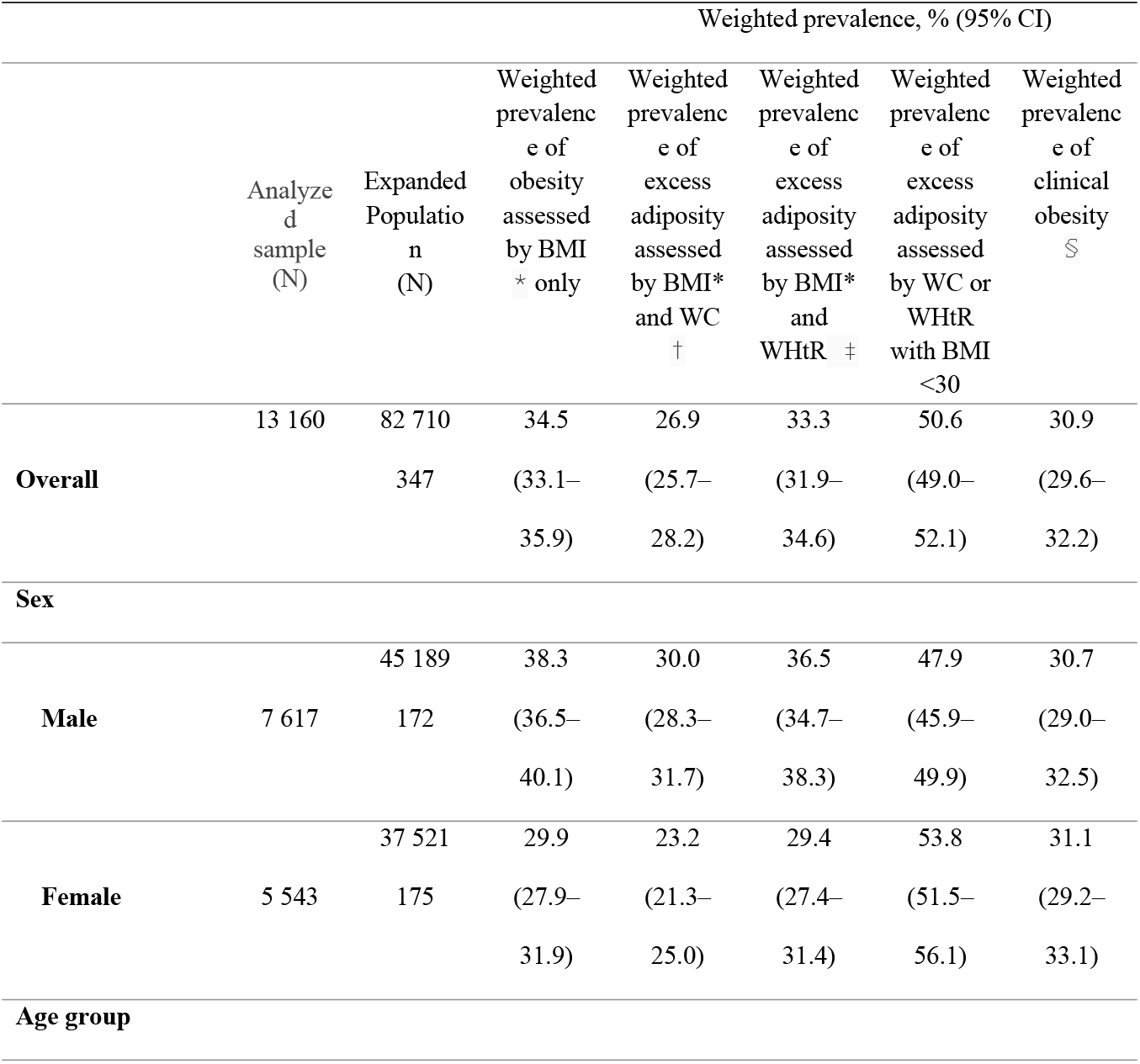
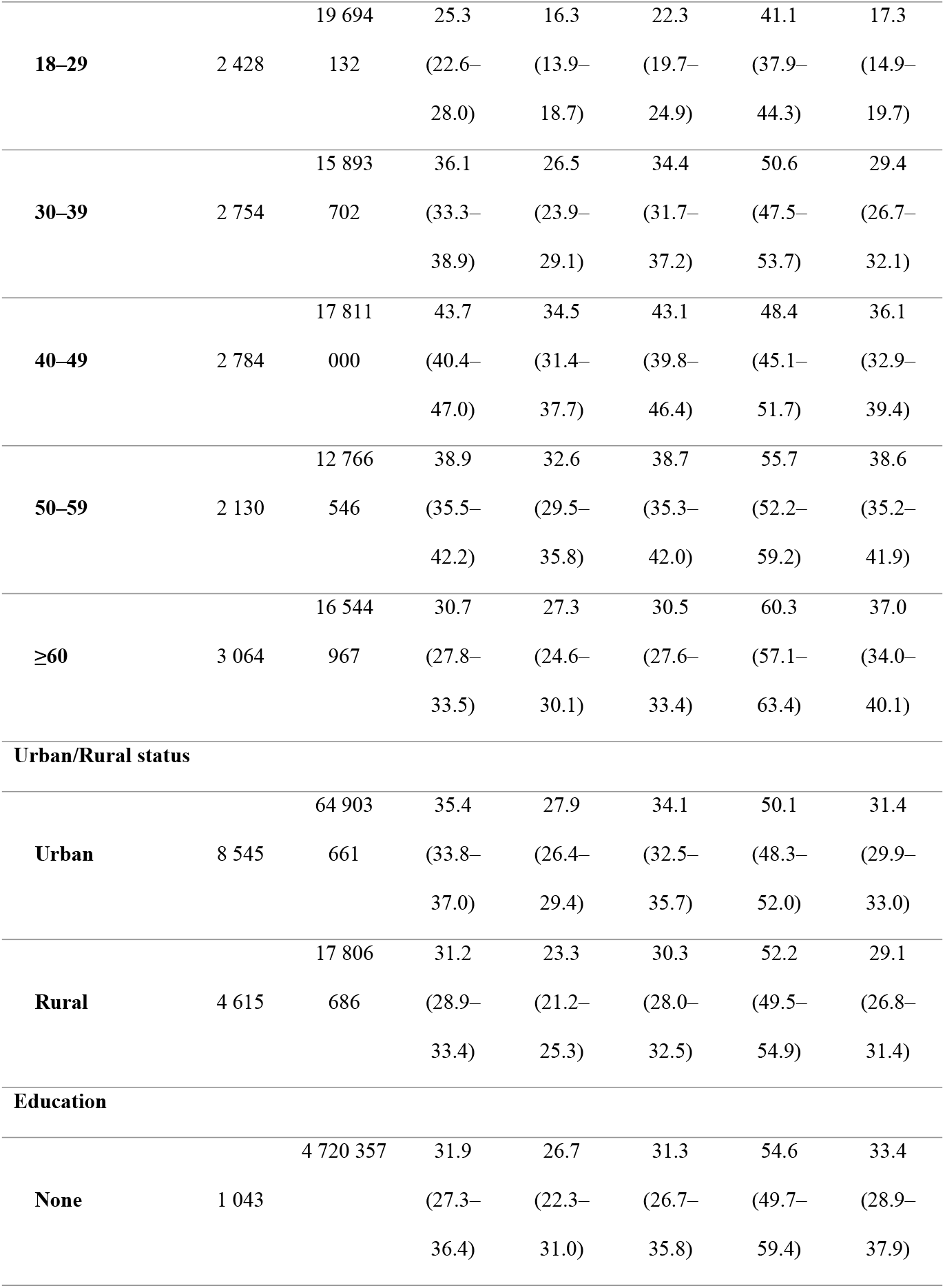
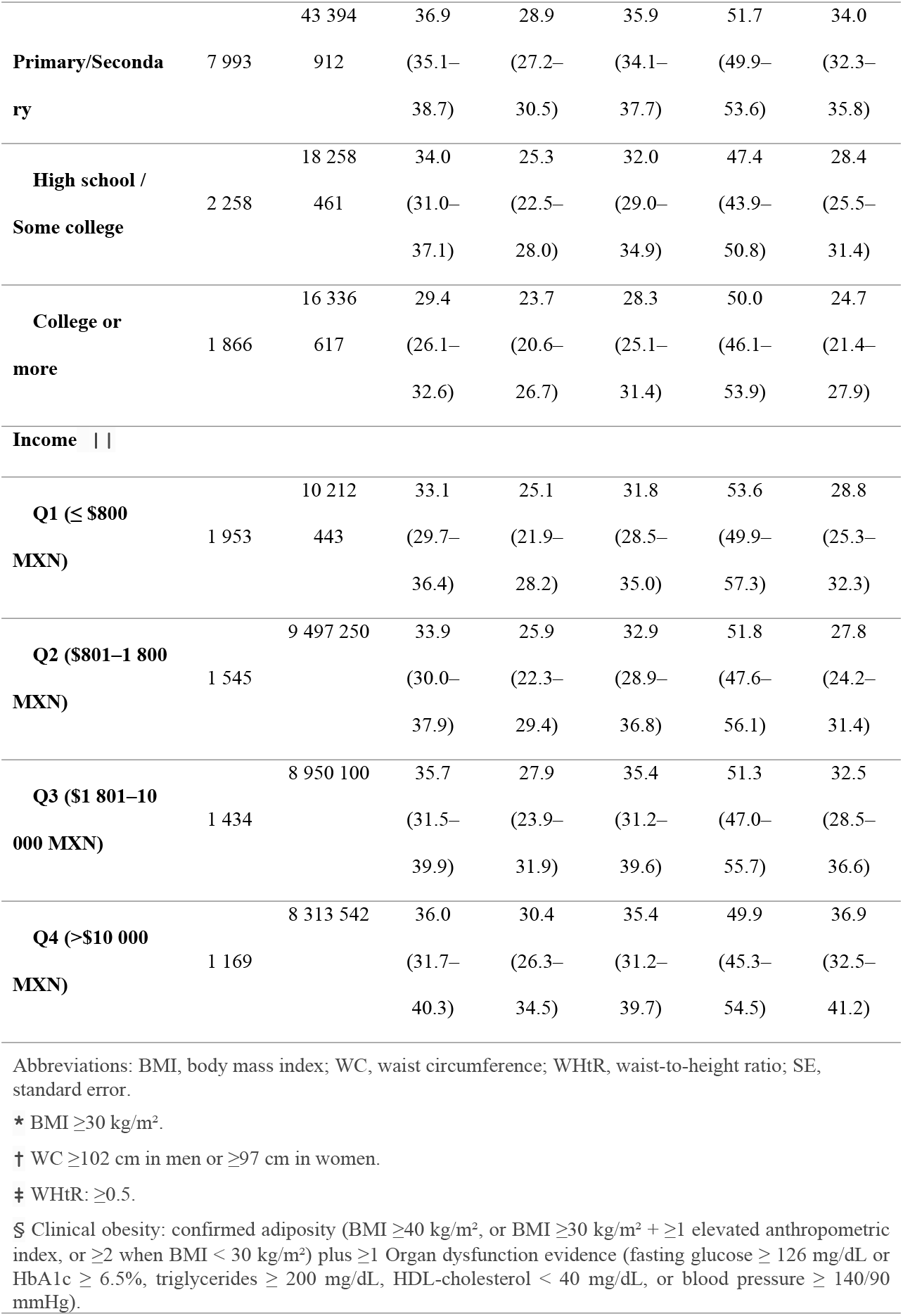
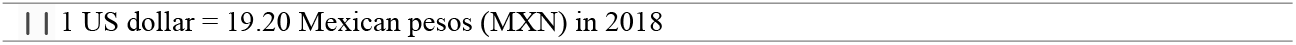
Weighted prevalence of excess adiposity and clinical obesity in Mexican adults.

Among adults with BMI <30 kg/m^2^, half (50.6%) showed signs of increased central adiposity, which was more common in women (53.8%) and older adults (60.3% among those aged ≥60 years). Clinical obesity was more prevalent in urban than rural settings (31.4% vs 29.1%). Prevalence was highest among adults with primary or secondary education (34.0%) and lowest among those with a college degree or higher (24.7%)

In adults with clinical obesity, 75% met criteria based on BMI ≥30 kg/m^2^ combined with at least one elevated anthropometric index, whereas 25% met criteria despite BMI <30 kg/m^2^ by having at least two elevated anthropometric indices (Figure). The proportion of clinical obesity with BMI <30 kg/m^2^ increased with age, from 12% among adults aged 18–29 years to 33% among those aged ≥60 years, and was similar between men and women (≈20%).

**Figure 1.**
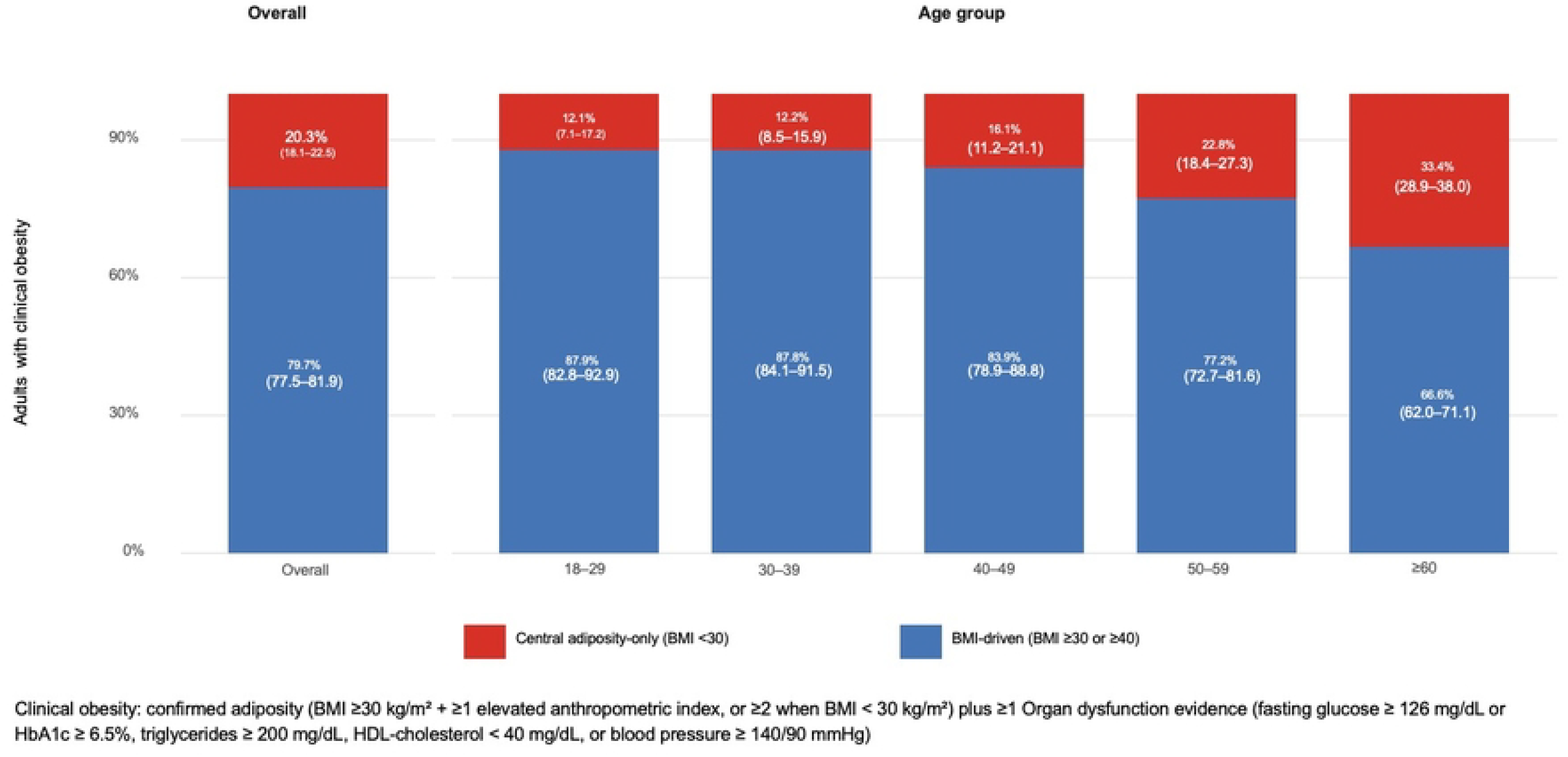
Clinical obesity: confirmed adiposity(BMI ≥ 30 kg/m^2^ + ≥ 1 elevated anthropometric index. or ≥ 2 when BMI < 30 kg/m^2^) plus ≥ 1 Organ dysfunction evidenoe(fasting glucose ≥ 126 mg/dL or HbAlc > 6.5%, triglycerides ≥ 200 mg/dl, HDL– Cholesterol< 40mg/dl, or blood pressure ≥ 140/90mmHg)

## DISCUSSION

Applying the Lancet Diabetes & Endocrinology Commission definition in Mexico uncovered underdiagnosis of clinical obesity when relying solely on BMI. Although the overall prevalence estimates were similar to those using combined anthropometric criteria, about one-quarter of clinical obesity cases occurred in adults with a BMI less than 30 kg/m^2^, especially among older individuals. These findings align with previous evidence from Hispanic and US populations. In the National Health and Nutrition Examination Survey, Aryee et al. demonstrated that 98–99% of adults with BMI ≥30 had confirmed excess adiposity, indicating that BMI rarely overestimates adiposity at higher levels but often underestimates it at lower levels.(7) Similarly, Palmer et al. found that 87.9% of Mexican Americans with a BMI under 30 kg/m^2^ had excess adiposity, as measured by dual-energy x-ray absorptiometry or waist circumference, a pattern reflected in our nationally representative data.(8)

Longitudinal data from Mexico support the clinical importance of this adiposity-only phenotype. In the Mexico City Prospective Study, Gnatiuc et al. showed that both general and abdominal adiposity were strongly linked to all-cause mortality, and that waist circumference and waist-to-hip ratio remained strong predictors of mortality even after adjusting for BMI. (9) These findings show that central adiposity poses a significant risk regardless of overall body size, highlighting the limitations of BMI as a standalone measure for risk assessment in Mexican adults.

Notably, this adiposity-only phenotype has been associated with significant clinical risks. Fourman et al. demonstrated that individuals with excess adiposity despite a BMI <30 kg/m^2^ had a higher risk of organ dysfunction and a greater occurrence of diabetes, cardiovascular events, and mortality.(10)

Although our analysis was limited by the lack of direct imaging-based measures of adiposity and incomplete characterization of organ dysfunction, these findings highlight the shortcomings of BMI as the sole metric. Incorporating waist-based indices into routine clinical assessments and population surveillance could enhance the identification of individuals with clinically significant obesity, particularly in populations like Mexico, where cardiometabolic risk remains high despite modest BMI levels.

## Data Availability

Underlying microdata from the 2018 to 2019 Mexican National Health and Nutrition Survey (ENSANUT) are third party data owned by the Instituto Nacional de Salud Pública (National Institute of Public Health of Mexico) and are publicly available without restriction at https://ensanut.insp.mx

## AUTHOR CONTRIBUTIONS

**MC-TC**: Conceptualization, Methodology, Data Curation, Formal Analysis, Writing - Original Draft; **NE-AV**: Data Curation, Formal Analysis, Writing - Review & Editing; **M-GA**: Writing - Review & Editing; **D-AG**: Writing - Review & Editing; **P-MA**: Conceptualization, Methodology, Writing - Review & Editing, Supervision.

## Acknowledgments

Data sharing statement. The data analyzed in this study are publicly available through the Instituto Nacional de Salud Pública (National Institute of Public Health of Mexico). The 2018 to 2019 ENSANUT dataset, including the data dictionary, can be accessed at https://ensanut.insp.mx without restriction. No new data were generated for this study.

